# Modelling the effect of a border closure between Switzerland and Italy on the spatiotemporal spread of COVID-19 in Switzerland

**DOI:** 10.1101/2021.05.19.21257329

**Authors:** Mathilde Grimée, Maria Bekker-Nielsen Dunbar, Felix Hofmann, Leonhard Held, on behalf of the SUSPend modelling consortium

## Abstract

We present an approach to extend the Endemic-Epidemic (EE) modelling framework for the analysis of infectious disease data. In its spatiotemporal application, spatial dependencies have originally been captured by a power law applied to static neighbourhood matrices. We propose to adjust these weight matrices over time to reflect changes in spatial connectivity between geographical units. We illustrate this extension by modelling the spread of coronavirus disease 2019 (COVID-19) between Swiss and bordering Italian regions in the first wave of the COVID-19 pandemic. We adjust the spatial weights with data describing the daily changes in population mobility patterns, and indicators of border closures describing the state of travel restrictions since the beginning of the pandemic. We use these time-dependent weights to fit an EE model to the region-stratified time series of new COVID-19 cases. We then adjust the weight matrices to reflect two counterfactual scenarios of border closures and draw counterfactual predictions based on these, to retrospectively assess the usefulness of border closures. We observed that predictions based on a scenario where no closure of the Swiss-Italian border occurred increased the number of cumulative cases in Switzerland by a factor of 2.5 over the study period. Conversely, a closure of the Swiss-Italian border two weeks earlier than implemented would have resulted in only a 12% decrease in the number of cases and merely delayed the epidemic spread by a couple weeks. Despite limitations in the current study, we believe it provides useful insight into modelling the effect of epidemic countermeasures on the spatiotemporal spread of COVID-19.

## 1. Introduction

The many types of uncertainty surrounding an ongoing emerging infectious disease outbreak with future endemic potential, such as coronavirus disease 2019 (COVID-19), motivates the use of statistical modelling to investigate current and future outbreaks of the disease. Not understanding the uncertainty of the true scale of the disease, e.g. through unreported cases, leads to difficulties in assessing the impact of required interventions [1, 2]. Multiple epidemic data sources provide valuable information on different aspects of such an outbreak, but require appropriate statistical techniques to incorporate the associated uncertainties.

One such statistical tool is the endemic-epidemic (EE) modelling framework, created for the analysis of infectious disease surveillance data [3]. It is a multivariate time series model that additively decomposes incidence into an endemic and an epidemic component. The epidemic component captures incidence driven by previous case counts, or force of infection, and the endemic component captures exogenous contributions to incidence (such as seasonal, socio-economic or demographic factors) [1, 3].

The best demonstration of the framework’s flexibility must be the wide range of infectious diseases it has been applied to. It can capture the incidence of infectious diseases of different types, with varying natural histories, vaccine preventability, and vector dynamics. In addition, the framework has already been widely used in analysis of COVID-19 outbreaks. Bekker-Nielsen Dunbar and Held [1] give an overview of the model’s various applications. This modelling approach can handle the heterogeneities and interdependencies in high-dimensional count time series and is useful for analysing infectious disease surveillance data which is stratified by geographic regions.

The EE framework was originally designed to fit weekly surveillance data. However, in light of the COVID-19 pandemic and the availability of daily case counts, the model has been extended to include higher-order lags. This means that the autoregressive process can be driven by a larger time-span of past case counts. In particular, this allows for the inclusion of infectiousness from the whole serial interval of COVID-19. Various epidemiological publications have used this feature in their analysis of COVID-19 incidence in Italy [4, 5], Germany [6], England [7], and the African continent [8] among others.

A key feature of the EE framework that will be discussed in the present paper is the possibility to include spatial dependencies, in addition to temporality. This allows for the modelling of transmission between regions over time. The spatial dependencies are included as weights in the autoregressive process of the epidemic component. The strength of spatial connectivity between regions, and the resulting transmissibility of the disease between them, can be formalised by the power law formulation introduced by Meyer and Held [9]. It reflects the decay of transmissibility between regions as the distance between them increases. Other groups have considered this approach in their endemic-epidemic modelling approaches [7, 8]. It has for instance been used by Ssentongo et al. [8] to model the effect of various socio-economic and meteorological factors, as well as containment strategies on the within- and between-country transmission in the African continent.

In this paper, we show that this approach can be extended with time-dependent weight matrices driving the epidemic component of the EE framework. We showcase how the static weights derived from the power law formulation can be adjusted over time to reflect the changing spatial relationship between geographical units as the mobility of the population shifts as a result of travel bans and border closures. This approach is of particular interest in the context of COVID-19, as in the first half of 2020 mobility-related social distancing interventions such as travel restrictions, border closures, and visit bans were imparted in many parts of the world, including within the Schengen area, in which the regions we consider here are contained. This was in spite of the World Health Organisation advising against the use of such measures at the time [10]. Mobility restrictions are only considered to work if protective sequestration is feasible, i.e. autochthonous transmission has not yet occurred in the territory and so only imported cases are of concern. In our work we adjust the weight matrices according to both within-region mobility (measured via smartphones) and between-region mobility (using information from the International Office for Migration). This provides a level of sophistication beyond simple mobility restriction/no mobility restriction indicators, as seen by e.g. Woo et al. [11] who used a similar model to investigate the spatiotemporal spread of COVID-19.

The possibility to adjust spatial weights over time gives us the opportunity to simulate different counter-factual scenarios, representing different timelines of pandemic countermeasures targeting population mobility. This in turn allows us to retrospectively evaluate the effect of border closures as a pandemic countermeasure. We can thereby provide evidence to improve public health response and aid in decisions on optimal infectious disease control strategies, particularly when to initiate travel restrictions.

## 2. Methods

### 2.1. Methodological framework

We examined the role of a border closure between Italy and Switzerland on the spatiotemporal spread of COVID-19 through the following steps:

1. Fit an EE model to the region-stratified time series of number of confirmed new COVID-19 cases from 24^th^ February 2020 until 4^th^ August 2020;
2. Retrospectively predict from the fitted model, given data until 2^nd^ March 2020, under scenarios:
  a. Closure of the Swiss-Italian border on 17^th^ March 2020, i.e. as really implemented (baseline scenario)
  b. No closure of the Swiss-Italian border (counterfactual scenario A)
  c. Closure of the Swiss-Italian border on 3^rd^ March 2020, two weeks earlier than in the baseline scenario (counterfactual scenario B);
3. Determine the total and region-specific number of cases in Switzerland based on the baseline and counterfactual scenarios from step 2; and
4. Examine the difference in total and region-specific incidence in Switzerland, to determine the usefulness of a border closure.

To fit our model, we included data from 24^th^ February 2020, as cases have been recorded in Switzerland and Italy after that date, until 4^th^ August 2020, as we only wanted to capture the dynamics of the first wave of the COVID-19 epidemic in Switzerland. The cutoff date of 2^nd^ March 2020 for the (counterfactual) predictions was chosen as the date after which the three scenarios might diverge.

Costantino et al. [12] consider similar scenarios when evaluating the effect of travel bans on COVID-19 spread in Australia.

### 2.2. Data

We were interested in analysing the COVID-19 spread in Switzerland and in the neighbouring regions in Italy. We considered data from the regions on the second level of the Nomenclature of Territorial Units for Statistics (NUTS-2). We included the seven Swiss regions, and the four bordering Italian regions, as listed in Table 1. A map of these eleven regions with their respective NUTS-2 code can be found in the supplementary materials.

**Table 1:** Regions included in the analysis

| Country | Regions (NUTS-2 code) |
| --- | --- |
| Switzerland | Région lémanique (CH01), Espace Mittelland (CH02), Nordwestschweiz (CH03), Zürich (CH04), Ostschweiz (CH05), Zentralschweiz (CH06), Ticino (CH07) |
| Italy | Piemonte (ITC1), Valle d’Aosta/Vallée d’Aoste (ITC2), Lombardia (ITC4), Provincia Autonoma di Bolzano/Bozen (ITH1) |

For each of these regions, we considered daily case data, population data, daily or weekly testing data, daily temperature data, data on public holidays, and data on border closures and population movement. Information on data sources and manipulations is available in the supplementary materials.

### 2.3. Model

We considered the EE modelling framework. Given daily disease surveillance case counts *Y*_*rt*_ indexed by NUTS-2 region (*r*) and day (*t*), the region-stratified EE model was given by [13, 14]

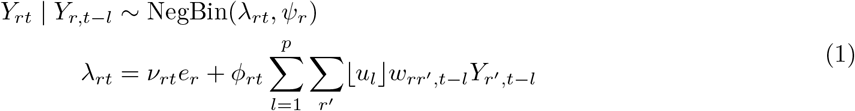

where *ψ*_*r*_ is a region-specific overdispersion parameter, *e*_*r*_ are population fractions, ⌊.⌋ denotes normalised weights, ⌊*u*_*l*_⌋ is the serial interval distribution, *w*_*rr′,t − l*_ is the transmission between regions *r* and *r*′ at day *t − l* (see Section 2.4), *p* is the maximum lag, and *ν*_*rt*_ and *ϕ*_*rt*_ are log-linear predictors. The *u*_*l*_ are shifted Poisson weights as recommended by Bracher and Held [14] for daily disease counts:

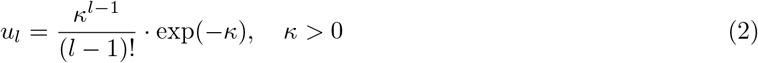

where *κ* is an unknown weighting parameter estimated from the data. We considered a maximum lag of *p* = 7

To determine optimal model specification, different model formulations were compared using model selection criteria. Here we used the Bayesian information criterion (BIC). In the final model, we included an indicator for weekends and country-specific public holidays, as medical facility clinic and laboratory operating hours may be reduced and population health-seeking activities may differ on weekends or public holidays compared with weekdays. We also included a country indicator, as we expected to see differences in endemicity between regions located in Switzerland and regions located in Italy. Additionally, we included indicators for each Swiss region bordering Italy (CH01, CH05, and CH07), as potential differences in underreporting between Italy and Switzerland would make it harder to capture the whole spatiotemporal spread by the power law alone. We also included country-specific (log-transformed) testing rates, as higher testing rates were expected to increase the number of cases. In the epidemic component, we further included a gravity model to reflect how more populated regions attract more imported cases [9, 15]. As a proxy for urbanisation, we included the log-population of the largest city in each region [16, 17]. We included log-temperature and yearly seasonality in our model, as we expected some form of seasonality in transmission related to seasonal changes in human behaviour. Finally, we included a simple linear time trend. All continuous covariates were centred.

The endemic predictor in our model was given by:

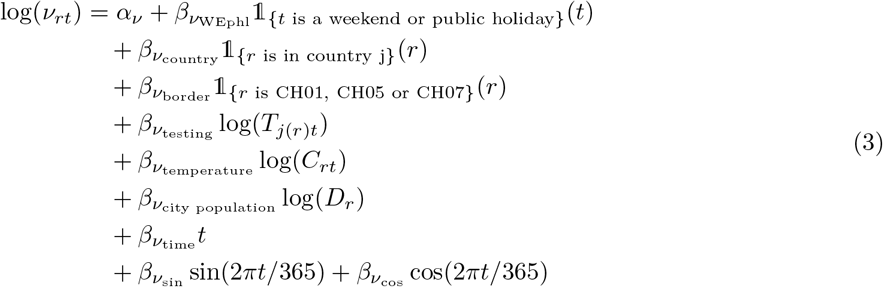

and the epidemic predictor was given by:

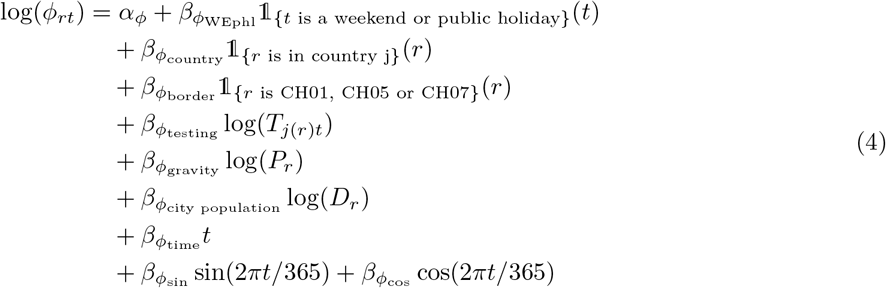

where *α* are overall intercepts, *T*_*j*(*r*)*t*_ is the country-specific testing rate at time *t, C*_*rt*_ is the region-specific temperature at time *t, D*_*r*_ is the population of the largest city in region *r, P*_*r*_ is the population of *r*, and *j*(*r*) describes the country in which region *r* is located and takes values “Switzerland” or “Italy”. The predictor 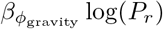 describes the gravity model [15].

### 2.4. Time-dependent matrix of neighbourhood order

To represent the spatial relationship between regions, driving the transmission between regions, as it changes over time a time-dependent matrix *W*_*t*_ of neighbourhood order was included in our model. We started from a static matrix of neighbourhood order *O* (see supplementary materials) and adjusted it to reflect the movement patterns and border closures between regions over time. We proceeded as follows:

1. To reflect the decreasing propensity of COVID-19 to spread between regions, as the distance between these regions increases, we implemented a power law model, as described in Meyer and Held [9]. Concretely, each entry of the matrix equals the inverse of the neighbourhood order, scaled by a decay parameter *d*. That is, the matrix entries are given by:

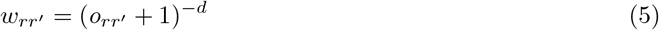

We set the parameter *d* = 1.6 [9]. This parameter is scale-free; it is independent of the units of distance [18]. It can thus be applied to neighbourhood order as a metric of distance. All further manipulations were not symmetric anymore, as movement between regions is not expected to be either. From here on, matrix rows represent “travel from” and columns represent “travel to”.
2. The Facebook^2^ Mobility data [19] allowed us to perform a daily adjustment of the matrix to reflect the change in population mobility. The time-dependent entry is given by:

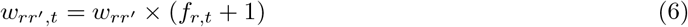

where *f*_*r,t*_ is the value of the Facebook movement change variable for region *r* at time *t* (see supplementary materials and [20]). The Facebook metric reports change in the quantity of movement from inhabitants of a region *r* and not change in movement direction. Therefore we could not directly infer the impact this change in movement has for the travel between *r* and each *r*′ individually. We assumed that it is uniform across all *r*′ and therefore adjusted whole matrix rows with the Facebook metric.
3. International Organisation of Migration (IOM) data allowed us to further adjust the time-dependent matrix to reflect border closures. The time-dependent entry is given by:

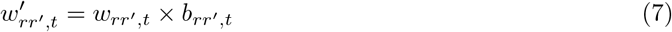

where *b*_*rr′*_,*t* is the state of restrictions to travel from region *r* to region *r*′, at time *t*. The IOM metric is qualitative and reports different border states as “No restrictions”, “Partial restrictions”, “Total restrictions”, and “No official restrictions reported”. We quantified these different border states as follows:

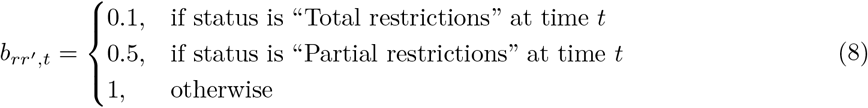

The time-dependent matrices are visualised in Figure 1 for the 1^st^ day of each month, after each step of adjustment.

**Figure 1:**
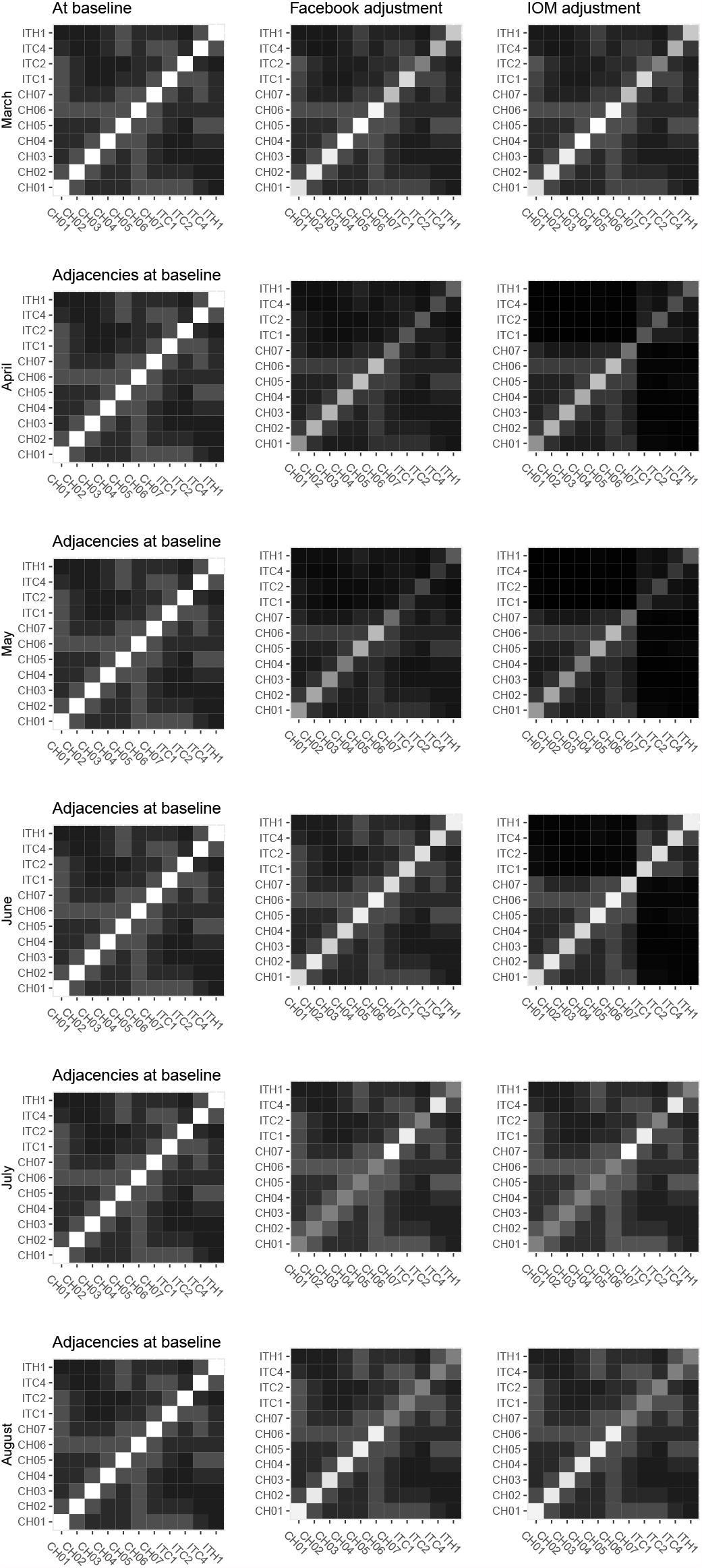
Time-dependent weight matrices after each step of adjustment

It should be noted that the ordering of regions in the matrices is arbitrary.

### 2.5. Counterfactual prediction of case counts

To examine the effect of border closures in place, we adjusted the neighbourhood matrices according to the three scenarios described under Section 2.1. For counterfactual scenario A, we implemented no border closures, which means that we only adjusted the matrix with the time-dependent Facebook metric *f*_*r,t*_. For counterfactual scenario B, we adjusted the matrix with *f*_*r,t*_ and, additionally to the border closures implemented in the baseline scenario, we implemented travel restrictions from Italy to Switzerland (*b*_*rr′*_,*t* = 0.1) between 3^rd^ March 2020 and 16^th^ March 2020.

We obtained parameter estimates by fitting the EE model described in Section 2.3 to the region-stratified time series of confirmed COVID-19 cases, based on the time-varying matrix of neighbourhood order from the baseline scenario. Then, given data from 24^th^ February 2020 to 2^nd^ March 2020, we predicted from the model, based on the weight matrices adjusted for counterfactual scenarios A and B. We assessed both predictions to determine the usefulness of border closures, reflected in both the total number of cases and the cases by region.

To obtain predictions based on our EE model, we used the predictive moments approach [13, 21]. To compare the predicted scenarios, we considered the relative increase between each counterfactual scenario (A and B) and the baseline scenario, and thus calculated

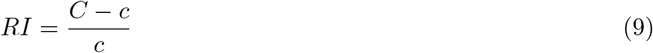

where *C* is the cumulative (total or region-specific) number of cases under the counterfactual scenario at hand, and *c* is the cumulative (total or region-specific) number of cases under the baseline scenario on 4^th^ August 2020 (end of the study period).

### 2.6. Implementation

This work used the EE modelling framework and associated `hhh4` package ecosystem, in particular the R (version 4.0.2) packages `surveillance` (version 1.19.1), and `hhh4addon` (version 0.0.0.0.9013).

## 3. Results

We fitted an EE model to the region-stratified time series of confirmed COVID-19 cases between 24^th^ February 2020 and 4^th^ August 2020 and the according covariates as described in Section 2.

It includes a weekend and (national) public holiday indicator, a country indicator, an indicator for the Swiss border regions, testing rates, temperature, population, population of the largest city, a time trend and seasonality. The coefficients of the model and the corresponding standard errors are shown in Table 2, and the fitted values are depicted in Figure 2. As is evident from Figure 2, the model fit depends on time. In particular in the first months, we see a slight underestimation of cases in most regions. It is interesting to note the different proportions of endemic to epidemic incidence in the different regions. We see the highest share of endemic incidence in Italy and Ticino (CH07). Given that the first COVID-19 cases in Switzerland were recorded in Ticino, and imported from Italy [22], this makes sense. The incidence in the French and German speaking regions of Switzerland can primarily be attributed to epidemic spread.

**Table 2:** Coefficient estimates and corresponding standard errors (SE) for the EE model given by (1), (2), (3), and (4)

|  | Estimate | SE |  | Estimate | SE |
| --- | --- | --- | --- | --- | --- |
| $\beta_{\phi_{\text{gravity}}}$ | 0.94 | 0.05 | $-\log(\psi_{\text{CH01}})$ | 2.34 | 0.18 |
| $\beta_{\phi_{\text{testing}}}$ | 0.34 | 0.05 | $-\log(\psi_{\text{CH02}})$ | 2.54 | 0.23 |
| $\beta_{\phi_{\text{time}}}$ | 0.02 | 0.01 | $-\log(\psi_{\text{CH03}})$ | 2.4 | 0.25 |
| $\beta_{\phi_{\text{WEphl}}}$ | -0.54 | 0.05 | $-\log(\psi_{\text{CH04}})$ | 1.46 | 0.2 |
| $\beta_{\phi_{\text{country}}}$ | 2.07 | 0.13 | $-\log(\psi_{\text{CH05}})$ | 1.57 | 0.21 |
| $\beta_{\phi_{\text{border}}}$ | 0.18 | 0.05 | $-\log(\psi_{\text{CH06}})$ | 2.2 | 0.26 |
| $\beta_{\phi_{\text{city population}}}$ | 0.36 | 0.04 | $-\log(\psi_{\text{CH07}})$ | 1.34 | 0.23 |
| $\beta_{\phi_{\text{sin}}}$ | -0.68 | 0.1 | $-\log(\psi_{\text{ITC1}})$ | 0.78 | 0.13 |
| $\beta_{\phi_{\text{cos}}}$ | 1.84 | 0.37 | $-\log(\psi_{\text{ITC2}})$ | 0.23 | 0.2 |
| $\alpha_{\phi}$ | -2.94 | 0.15 | $-\log(\psi_{\text{ITC4}})$ | 1.45 | 0.12 |
| $\beta_{\nu_{\text{testing}}}$ | 1.06 | 0.12 | $-\log(\psi_{\text{ITH1}})$ | 0.47 | 0.19 |
| $\beta_{\nu_{\text{time}}}$ | 0.18 | 0.03 | | | |
| $\beta_{\nu_{\text{WEphl}}}$ | 0.35 | 0.13 | | | |
| $\beta_{\nu_{\text{country}}}$ | -1.45 | 0.59 | | | |
| $\beta_{\nu_{\text{border}}}$ | 2.84 | 0.5 | | | |
| $\beta_{\nu_{\text{temperature}}}$ | 0.04 | 0.01 | | | |
| $\beta_{\nu_{\text{city population}}}$ | 0.89 | 0.05 | | | |
| $\beta_{\nu_{\text{sin}}}$ | -4.88 | 0.61 | | | |
| $\beta_{\nu_{\text{cos}}}$ | 15.86 | 2.39 | | | |
| $\alpha_{\nu}$ | 1.48 | 0.32 | | | |

**Figure 2:**
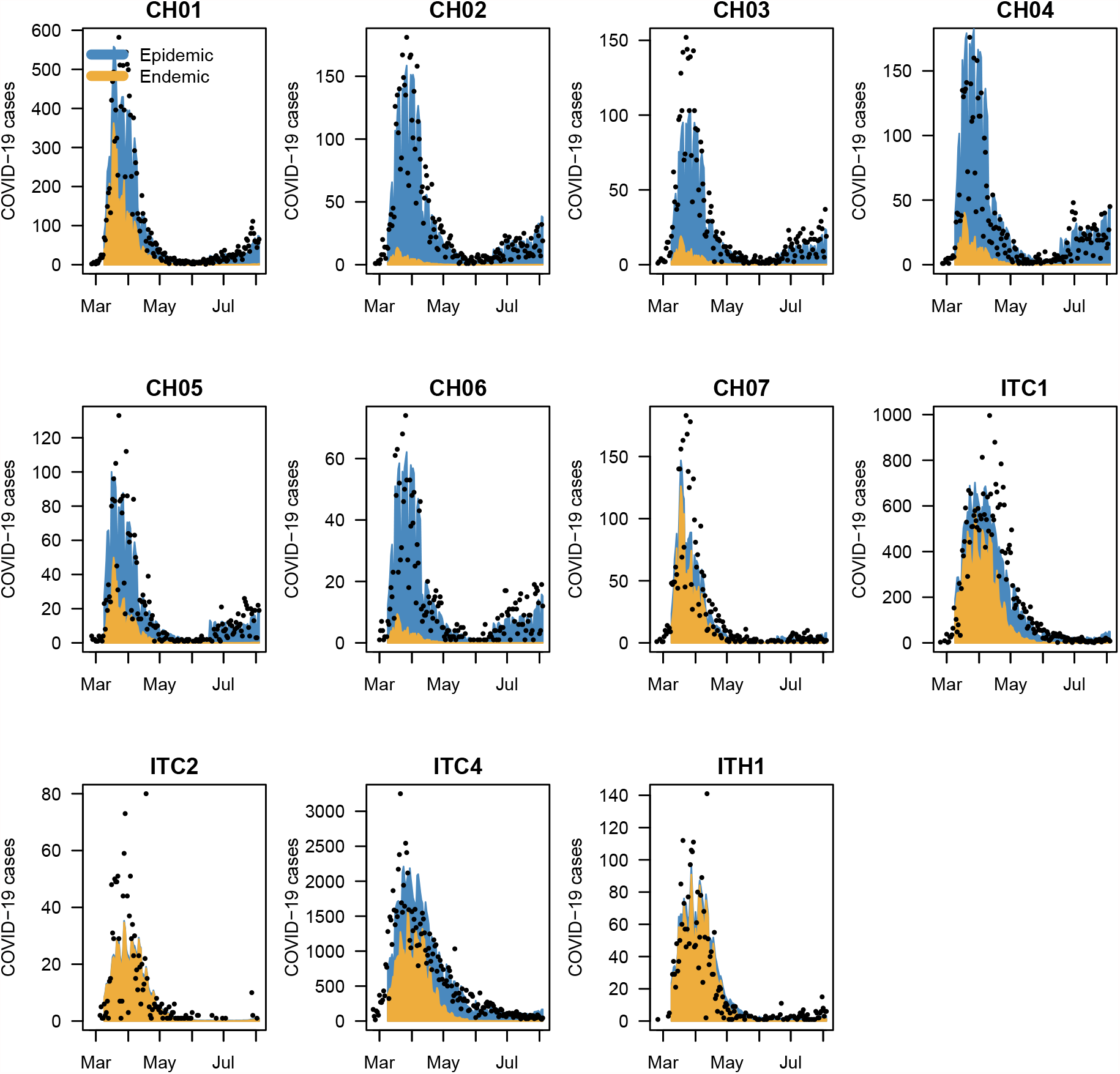
Fitted values for the model given by (1), (2), (3), and (4)

The estimated lag distribution can be found in Table 3, and the corresponding plot in the supplementary materials. Our findings are consistent with the estimates obtained by Ssentongo et al. [8].

**Table 3:** estimated lag distribution for the EE model given by (1), (2), (3), and (4)

| 1 | 2 | 3 | 4 | 5 | 6 | 7 |
| --- | --- | --- | --- | --- | --- | --- |
| 0.06 | 0.16 | 0.24 | 0.23 | 0.17 | 0.10 | 0.05 |

Given this model, data from the 8 days between 24^th^ February 2020 and 2^nd^ March 2020, and the counterfactual weight matrices described in Section 2.5, we predicted total and region-specific cumulative cases until 4^th^ August 2020 under each scenario. The results are summarised in Table 4. To illustrate prediction accuracy, we plot a comparison of the daily incidence predicted under the baseline scenario, and observed incidence over the study period in the supplementary materials. Plots of the daily difference and ratio of new and cumulative cases between the counterfactual scenarios and the baseline scenario are available in the supplementary materials for the complete study period.

**Table 4:** Relative increase in cases for the counterfactual scenarios compared to baseline.

| Territory | Baseline scenario | Scenario A | Scenario B | Relative increase (A) | Relative increase (B) |
| --- | --- | --- | --- | --- | --- |
| Total | 169014.9 | 269082.0 | 164715.8 | 0.6 | 0.0 |
| CH | 35300.3 | 90008.5 | 31178.8 | 1.5 | -0.1 |
| IT | 133714.6 | 179073.5 | 133537.0 | 0.3 | 0.0 |
| CH01 | 13524.4 | 28049.9 | 12466.6 | 1.1 | -0.1 |
| CH02 | 4699.5 | 13750.7 | 3973.9 | 1.9 | -0.2 |
| CH03 | 2959.2 | 8612.3 | 2501.1 | 1.9 | -0.2 |
| CH04 | 6349.0 | 19444.2 | 5317.5 | 2.1 | -0.2 |
| CH05 | 2926.3 | 9014.4 | 2522.1 | 2.1 | -0.1 |
| CH06 | 1946.2 | 5882.1 | 1649.9 | 2.0 | -0.2 |
| CH07 | 2895.6 | 5254.9 | 2747.6 | 0.8 | -0.1 |
| ITC1 | 29736.0 | 39879.2 | 29692.6 | 0.3 | 0.0 |
| ITC2 | 1093.5 | 1173.3 | 1093.1 | 0.1 | 0.0 |
| ITC4 | 99692.8 | 134363.0 | 99561.2 | 0.3 | 0.0 |
| ITH1 | 3192.2 | 3658.1 | 3190.1 | 0.1 | 0.0 |

Under counterfactual scenario A, i.e. no border closure, we see a 1.6-fold increase in cases compared to baseline, with most excess cases happening in the first months. The peak number of excess cases compared to baseline would have been attained in early April with around 2000 additional daily cases (see supplementary materials for details). Overall, this scenario would have caused 100067 extra cases between 3^rd^ March 2020 and 4^th^ August 2020, with 54708 cases in Switzerland and 45359 in the bordering Italian regions.

Under counterfactual scenario B, i.e. earlier border closure, we see only a very small relative decrease overall. This would primarily have affected the early months, with a peak reduction of daily cases (c.a. − 450) around mid-March. From April onwards, no difference in daily cases would have been observed between scenario B and baseline (see supplementary materials for details). As we only implemented an internal border closure in Switzerland, i.e. only restricting entry in Switzerland from Italy, this decrease in cases naturally only affects the spread in Switzerland. Overall, this scenario would have prevented 4299 cases between 3^rd^ March 2020 and 4^th^ August 2020, of which the vast majority (4122) in Switzerland.

Figure 3 shows a timeline of predicted cases under each scenario for each of the regions. It is apparent that scenario A (blue) would have cause a marked increase in daily cases throughout the study period in all Swiss regions. The marginal effect of an earlier closure of borders (scenario B, yellow) can be clearly seen from the plot, as it remains barely distinguishable from the baseline scenario (pink) throughout the first wave.

**Figure 3:**
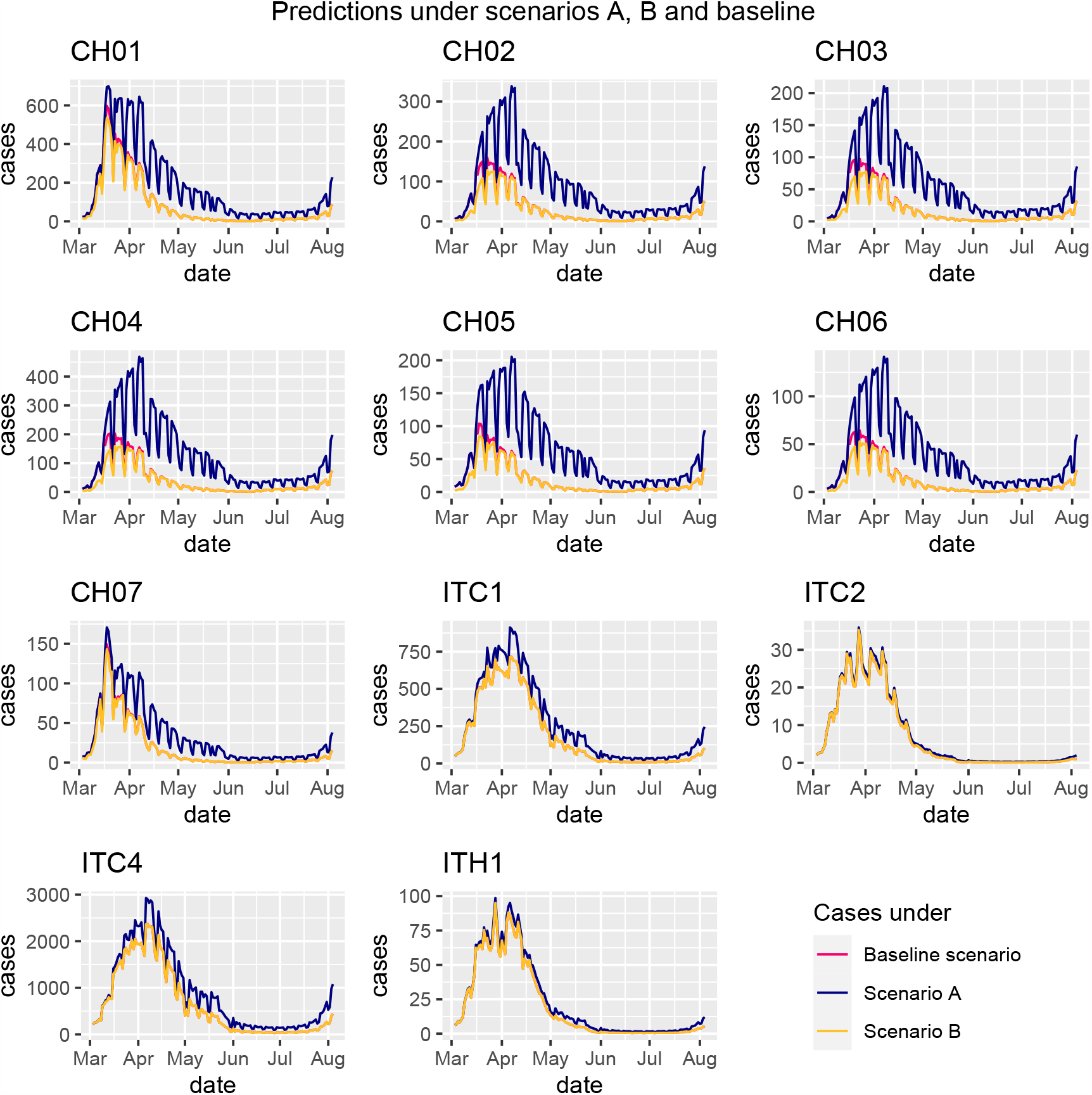
Predictions under scenarios A, B, and baseline

The results seem to support the baseline border closure scenario. Without the measures implemented, we would expect to have seen a tremendous rise in cases, whereas closing the border closures earlier would have merely delayed the spread of infection in Switzerland, and would overall have prevented only around 4300 cases.

## 4. Discussion

We highlighted the use of time-dependent weight matrices reflecting changes in spatial connectivity, to extend the traditional spatiotemporal EE framework. It is a particularly useful approach in the context of COVID-19, where pandemic countermeasures have targeted the spatial relations between geographical units with the implementation of border closures and travel restrictions, and where many data sources on patterns of population mobility have become available.

Some limitations in the present paper need to be addressed to draw unbiased conclusions from our results. The foundation of the spatial relationship between geographical units in this study is the power law application, upon which all further adjustments rely. It is therefore crucial that it reflects the spatial connectivity correctly. In this context, it would have been important that the corresponding decay parameter *d* be estimated from the data [13]. However, this parameter estimation is not (yet) compatible with the R-implementation of the higher-order lags *u*_*l*_, which are also crucial in the study of COVID-19. In this study, we chose to use a fixed parameter motivated by existing studies [9, 18], but we recognise that this could have weakened our time-dependent weights. Further work could examine how to facilitate harmonising the two approaches. Similarly, the implementation of the IOM metric in this study might need appropriate modification. As mentioned in the methods, the IOM metric is qualitative, and the way we chose to quantify the indicators could be disputed. Indeed, we have chosen to weigh “total restrictions” as 0.1, meaning that the strength of the spatial relationship between regions separated by a border would be weakened by a factor of 10 upon closure of the latter. Whether this accurately reflects reality remains to be determined.

It is also crucial that we address model fit, which naturally influences the subsequent predictions. In initial fittings of the EE model, we observed a particular underestimation of cases in the first wave for the regions CH01, CH05 and CH07, the three regions bordering Italy. Ideally, region-specific intercepts would have been implemented to address this, but this led to model divergence and thus unreliable parameter estimations. In this paper, we chose to address this by fitting a model with region-specific overdispersion, which significantly improved overall fit, and a specific indicator for the border regions, which improved fit in those regions. However, in the present formulation of the model, we still see an underestimation of cases in most regions in the first months. This leads us to conclude that there is more spatiotemporal spread happening than what can be captured by the power law alone. We suspect that this is due to differing levels of underreporting, particularly in the first wave, between Italy and Switzerland but also between individual regions. In light of these limitations, we recommend that further studies take underreporting and underascertainment of case counts into consideration. Simple multiplication factors can be used [23] as a first-order approach.

Moreover, a full picture of the border situation of Switzerland would be beneficial to gain precision, as the spatiotemporal spread of COVID-19 in Switzerland arguably can not only be explained by importation of cases from Italy and endemic factors. We intend to extend our work to additionally take the neighbouring regions in France, Germany, and Austria into consideration. Finally, the EE framework can be adjusted to incorporate pharmaceutical countermeasures such as vaccines [24]; an obvious direction to pursue were a similar analysis to be conducted for 2021.

All in all, we do believe that the present paper, extending the EE modelling approach with time-dependent spatial weights, provides useful insight into the analysis of the spatiotemporal spread of COVID-19 and can be an important stepping stone for further research informing decision-making around COVID-19 counter-measures.

## 5. Conclusion

We used time-dependent matrices of neighbourhood order between regions, adjusted through power laws, mobility data, and border status indicators, allowing us to explore spatial interventions’ impact on early cases of COVID-19 in Switzerland and Italy. This approach has allowed us to draw counterfactual predictions based on weight matrices adjusted for counterfactual scenarios of border closures. We have observed that predictions based on counterfactual scenario A, where no closure of the Swiss-Italian border occurred over the study period, would have almost doubled the cumulative number of cases over the study period. Additionally, based on counterfactual scenario B, where closure of the Swiss-Italian border occurred two weeks earlier than implemented in reality, predictions showed only marginally reduced numbers of cases and an epidemic spread merely delayed by a couple weeks, which is in line with the WHO’s statement [10].

We are unable to determine what an acceptable increase in cases would be as we are not policy makers, and as a result have not considered cut-off criteria or hypothesis tests. However, the total number of cases in the first wave of the pandemic almost doubled under the scenario where no border closures occurred.

## Data Availability

The code and data used for this article are available on the GitLab repository linked below.
Further description of the data availability is available in the supplementary materials.

https://gitlab.switch.ch/suspend/COVID-19-border-CH/

## Acknowledgements

This work would not be possible without the financial support of the Swiss National Science Foundation (https://data.snf.ch/covid-19/snsf/196247). They did not play any role in the formulation of this manuscript and the views expressed in this manuscript are those of the authors. The additional members of the SUSPend modelling consortium (in anti-alphabetical order by surname) are: Jon Wakefield, Sebastian Meyer, Niel Hens, Deborah Chiavi, and Johannes Bracher (https://suspend.pages.switch.ch/project). Some of the authors are residents of the regions considered in this analysis. The authors declare no further conflicts of interest.

## Credit taxonomy

- Mathilde Grimée: methodology, software, formal analysis, visualisation, and writing
- Maria Becker-Nielsen Dunbar: conceptualisation, methodology, software, funding acquisition, writing, project administration, and supervision
- Felix Hofmann: methodology, software, data curation, and visualisation
- Leonhard Held: conceptualisation, methodology, funding acquisition, writing, project administration, and supervision

## Footnotes

2 Our use of data provided by Facebook is not to be seen as an endorsement of Facebook as a company.

## References

[1] M. Bekker-Nielsen Dunbar and L. Held. Endemic-Epidemic framework used in COVID-19 modelling. RevStat, 18(5), 2020.

[2] P. J. Birrell, L. Wernisch, B. D. M. Tom, Held L., G. O. Roberts, R. G. Pebody, and D. De Angelis. Efficient real-time monitoring of an emerging influenza pandemic: How feasible? Ann. Appl. Stat, 14 (1):74–93, 2020. doi: 10.1214/19-AOAS1278.

[3] L. Held, M. Höhle, and M. Hofmann. A statistical framework for the analysis of multivariate infectious disease surveillance counts. Stat. Model, 36(22):187–199, 2005. doi: 10.1191/1471082X05st098oa.

[4] M. M. Dickson, G. Espa, D. Giuliani, F. Santi, and L. Savadori. Assessing the effect of containment measures on the spatio-temporal dynamic of COVID-19 in Italy. Nonlinear Dyn., 2020. doi: 10.1007/s11071-020-05853-7.

[5] D. Giuliani, M.M. Dickson, G. Espa, and F. Santi. Modelling and predicting the spatio-temporal spread of coronavirus disease 2019 (COVID-19) in Italy. BMC Infect. Dis, 20:700, 2020. doi: 10.1186/s12879-020-05415-7.

[6] C. Fritz and G. Kauermann. On the interplay of regional mobility, social connectedness, and the spread of COVID-19 in Germany. arXiv, 20, 2020. URL https://arxiv.org/abs/2008.03013.

[7] C. Fronterre, J. M. Read, B. Rowlingson, J. Bridgen, S. Alderton, P. J. Diggle, and C. P Jewell. COVID-19 in England: spatial patterns and regional outbreaks. medRxiv, 2020. doi: 10.1101/2020.05.15.20102715.

[8] P. Ssentongo, C. Fronterre, A. Geronimo, S. J. Greybush, P. K. Mbabazi, J. Muvawala, S. B. Nahalamba, P. O. Omadi, B. T. Opar, S. A. Sinnar, Y. Wang, A. J. Whalen, L. Held, C. Jewell, A. J. B. Muwanguzi, H. Greatrex, M. M. Norton, P. Diggle, and S. J. Schiff. Tracking and predicting the African COVID-19 pandemic. medRxiv, 2020. doi: 10.1101/2020.11.13.20231241.

[9] S. Meyer and L. Held. Power-law models for infectious disease spread. Ann. Appl. Stat, 8:1612–1639, 2014. doi: 10.1214/14-AOAS743.

[10] World Health Organization. Updated WHO recommendations for international traffic in relation to COVID-19 outbreak (29 February 2020), 2020. URL https://www.who.int/news-room/articles-detail/updated-who-recommendations-for-international-traffic-in-relation-to-covid-19-outbreak.

[11] H. Woo, O. Kwon, and J.-S. Yang. Spatiotemporal spread of infectious diseases and intra-or intertransmission: A gravity model approach. SSRN, 3731170, 2020. doi: 10.2139/ssrn.3731170.

[12] V. Costantino, D. J. Heslop, and C. R. MacIntyre. The effectiveness of full and partial travel bans against COVID-19 spread in Australia for travellers from China during and after the epidemic peak in China. J. Travel Med, 27(5), 2020. doi: 10.1093/jtm/taaa081.

[13] L. Held, S. Meyer, and J. Bracher. Probabilistic forecasting in infectious disease epidemiology: the 13th Armitage lecture. Stat. Med, 36(22):3443–3460, 2017. doi: 10.1002/sim.7363.

[14] J. Bracher and L. Held. Endemic-epidemic models with discrete-time serial interval distributions for infectious disease prediction. Int. J. Forecast, 2020. URL https://arxiv.org/abs/1901.03090.

[15] Y. Xia, O.N. Bjørnstad, B.T. Grenfell, and D.L. DeAngelis. Measles metapopulation dynamics: A gravity model for epidemiological coupling and dynamics. the american naturalist, 164(2):267––281, 2004.

[16] K. Kafadar and J. W. Tukey. U.S. cancer death rates: A simple adjustment for urbanization. Int. Stat. Rev, 61:257–281, 1993. doi: 10.2307/1403628.

[17] L. Knorr-Held and J. Besag. Modelling the risk of a disease in time and space. Stat. Med, 17:2045–2060, 1998. doi: 10.1002/(SICI)1097-0258(19980930)17:18<2045::AID-SIM943>3.0.CO;2-P.

[18] D. Brockmann, L. Hufnagel, and T. Geisel. The scaling laws of human travel. Nature, 2006. doi: 10.1038/nature04292.

[19] Facebook. Facebook data for good. movement range data, 2020. URL https://data.humdata.org/ dataset/movement-range-maps.

[20] Facebook Data for Good. Protecting privacy in Facebook mobility data during the COVID-19 response, 2020. URL https://research.fb.com/blog/2020/06/protecting-privacy-in-facebook-mobility-data-during-the-covid-19-response.

[21] J. Bracher and L. Held. A marginal moment matching approach for fitting endemic-epidemic models to underreported disease surveillance counts. Biometrics, pages 1–13, 2020. doi: 10.1111/biom.13371.

[22] Bundesamt für Gesundheit BAG. Neues coronavirus COVID-19: Erster bestätigter Fall in der Schweiz, 2020. URL https://www.bag.admin.ch/bag/de/home/das-bag/aktuell/medienmitteilungen.msg-id-78233.html.

[23] A. Noufaily. Underreporting and reporting delays. In Handbook of Infectious Disease Data Analysis, chapter 22, pages 437–454. Chapman & Hall/CRC Handbooks of Modern Statistical Methods, 2020.

[24] S. A. Herzog, M. Paul, and L. Held. Heterogeneity in vaccination coverage explains the size and occurrence of measles epidemics in German surveillance data. Epidemiol. Infect, 139(4):505–515, 2011. doi: 10.1017/S0950268810001664.

