## Supplementary materials for "Modelling the effect of a border closure between Switzerland and Italy on the spatiotemporal spread of COVID-19 in Switzerland"

[a] *Institute for Medical Information Processing, Biometry, and Epidemiology, Ludwig-Maximilians-Universität München*
[b] *Epidemiology, Biostatistics and Prevention Institute, University of Zurich, Hirschengraben 84, 8001 Zurich, Switzerland*
[c] *Swiss School of Public Health*
[d] *Life Science Zurich Graduate School*
[e] *Center for Reproducible Science, University of Zurich*

### Abstract

This document provides supporting information for "Modelling the effect of a border closure between Switzerland and Italy on the spatiotemporal spread of COVID-19 in Switzerland". In particular, data sources and manipulations are recorded, as well as more supporting plots for the results section.

### 1. Data sources and manipulations

In this section, we provide additional information on the data used in this analysis.

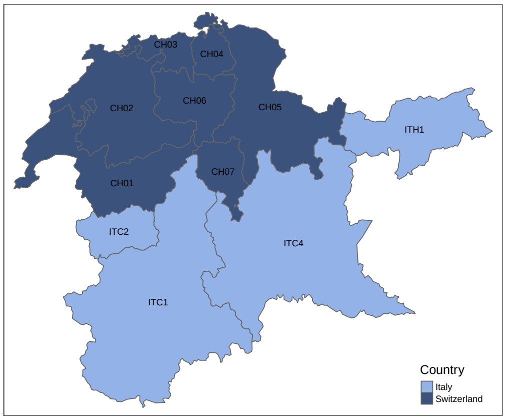


Figure 1: Map of the regions included in the analysis

#### *1.1. Case data*

Case data for Switzerland is provided by the Bundesamt für Gesundheit (BAG) [1]. The time series starts at 2020-02-24 and is updated on weekdays. However, all cases and deaths occurring on weekends are allocated to the respective weekend days and published the following Monday. The data is available at NUTS-3 level and aggregated to the NUTS-2 level. For Italy, case data is provided by the Presidenza del Consiglio dei Ministiri – Dipartimento della Protezione Civile (PCM-DPC) [2]. The data is available at the NUTS-2 level. The time series starts at 2020-02-24 and is updated daily. For the considered regions and time-frame, we

* Corresponding author
1 These authors contributed equally

observed two instances of negative numbers of new cases, which is impossible. However, as these barely deviated from 0 (one instance recorded -1 and another -10 new cases in a given region on a given day), we set new case counts on these days in these regions to zero.

#### *1.2. Population and popublation density data*

All population data was considered for the year 2020. It is available via Eurostat [3]. Data for the population of Swiss cities was retrieved from Swiss Federal Statistical Office [4]. Data for the population of Italian cities was retrieved from Istituto Nationale di Statistica [5].

#### *1.3. Data on public holidays.*

Information about national public holidays during the study period in Switzerland was retrieved from Eidgenössisches Justiz- und Polizeidepartement EJPD [6]. Information about national public holidays during the study period in Italy was retrieved from Governo Italiano Presidenza del Consiglio dei Ministri [7]. The public holidays used are summarised in Table 1:

| Country | Date | Holiday |
|---|---|---|
| Switzerland | 2020-04-10 | Good Friday |
| Switzerland | 2020-04-13 | Easter Monday |
| Switzerland | 2020-05-21 | Ascension day |
| Switzerland | 2020-06-01 | Whit Monday |
| Italy | 2020-04-13 | Easter Monday |
| Italy | 2020-05-01 | Labor Day |
| Italy | 2020-06-02 | Republic Day |

Table 1: National public holidays in Switzerland and Italy between 2020-02-24 and 2020-08-04

Public holidays that fall on a Saturday or Sunday are not recorded in Table 1.

#### *1.4. Testing data*

For Switzerland, testing data is found via the BAG [8]. The time series is available daily and goes from 2020-01-24 until the present. For Italy, testing data is obtained via ECDC [9]. The time series is available weekly and starts at week 5 of 2020 (week commencing 2020-01-27). Both datasets provide testing rates as number of tests per 100 000 inhabitants.

#### *1.5. Temperature data*

Temperature data was obtained from European Climate Assessment and Dataset [10]. This dataset contains daily temperature data for various weather stations. We matched each of the 11 regions in our analysis to the weather station closest to the centroid of the region, that did not contain missing data.

#### *1.6. Border closures*

Data on border closures was scraped from the web page of the International Organisation for Migration (IOM) [11]. This web page contains information on travel restrictions in matrix format, where the rows represent the countries that are imposing the restrictions and the columns are the countries to which restrictions are imposed upon. The data is available at regular intervals for both countries considered in our analysis, from 2020-03-08 and is updated regularly. In these matrices, border closures are recorded at levels “No restrictions”, “Partial restrictions”, “Total restrictions” and “No official restrictions reported”.

#### 1.7. Mobility data

To include the change in movement of populations in our model, we considered the Facebook[2] Data for Good dataset on movement range [12]. It contains data at NUTS-2 level for Italy, which can be used as is. The Swiss data is available at NUTS-3 level, which we can easily aggregate as a weighted average over the NUTS-2 region (with the weights being the population in the NUTS-3 regions). The variable of interest in that data set is `all_day_bing_tiles_visited_relative_change`. This variable is computed as the difference between movement on a given day, and movement at baseline, with respect to movement at baseline [13]. Movement at baseline is defined as the movement in the month of February 2020. It is recorded from 2020-03-01 and updated regularly.

#### 1.8. Matrix of neighbourhood order

We used a matrix of neighbourhood order $O$ in our model that describes the neighbourhood orders between the analysed regions. The data used to create this matrix was obtained from Eurostat [14]. The matrix $O$ is an $R \times R$ matrix, with $R$ being the number of analysed regions. The matrix entry $o_{rr'}$ equals the minimum of border crossings on the path between the two regions (e.g. $o_{rr'} = 1$ if $r$ and $r'$ are direct neighbours) [15]. This matrix is symmetric, i.e. $o_{rr'} = o_{r'r}$.

### 2. Relative increase ($\boldsymbol{RI}$) time series

In this section, we plot the difference and relative increase in new and cumulative cases over time between the counterfactual scenarios A and B, and the baseline scenario.

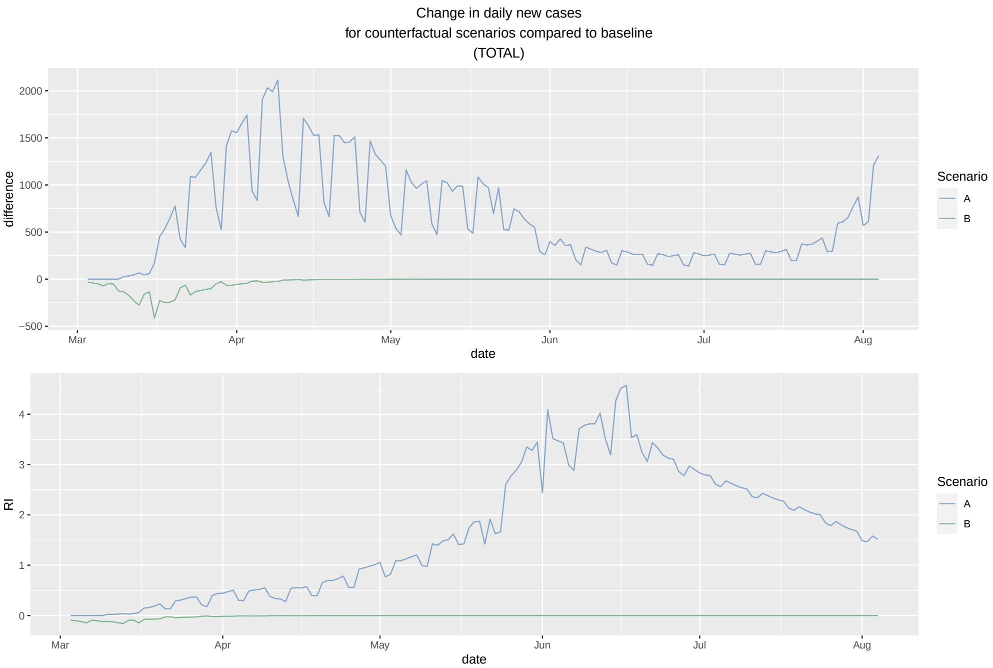


Figure 2: Change in daily new cases compared to baseline between 2020-03-02 and 2020-08-04

[2]Our use of data provided by Facebook is not to be seen as an endorsement of Facebook as a company.

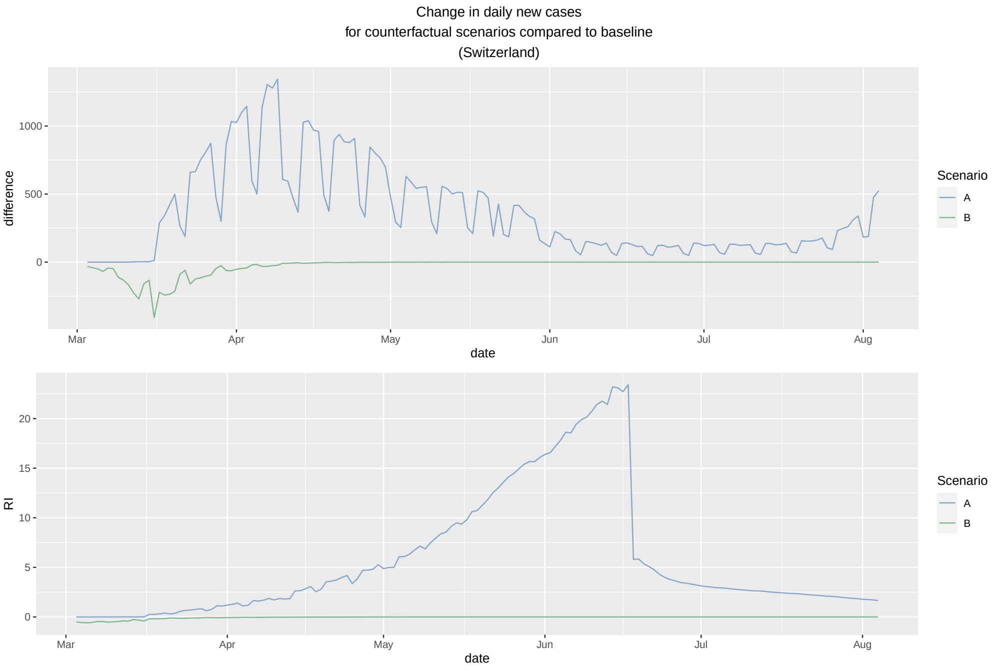


Figure 3: Change in daily new cases compared to baseline in Switzerland between 2020-03-02 and 2020-08-04

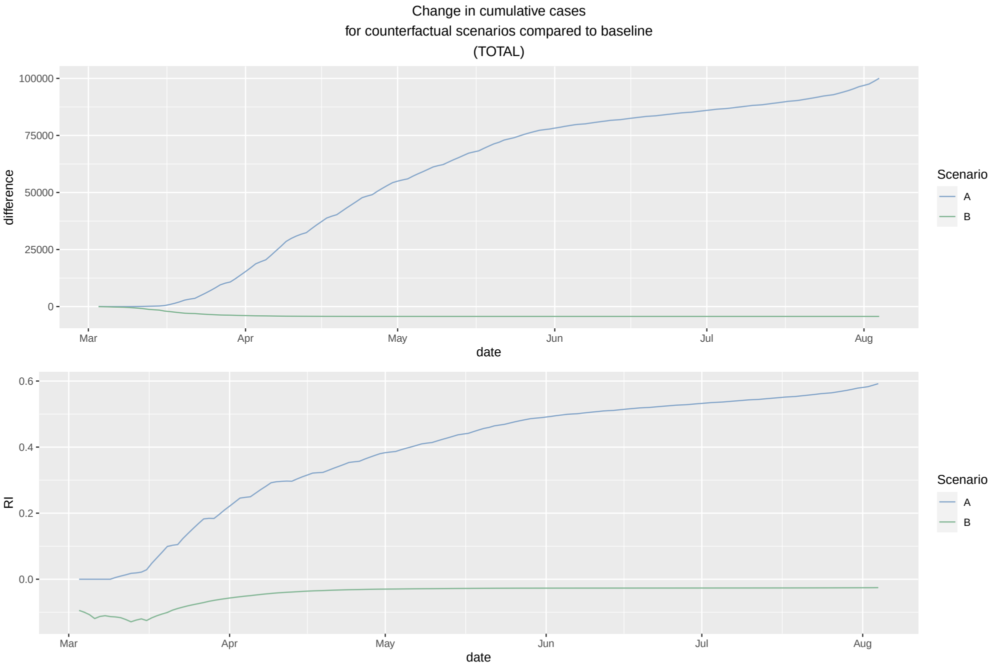


Figure 4: Total change in cumulative cases compared to baseline between 2020-03-02 and 2020-08-04

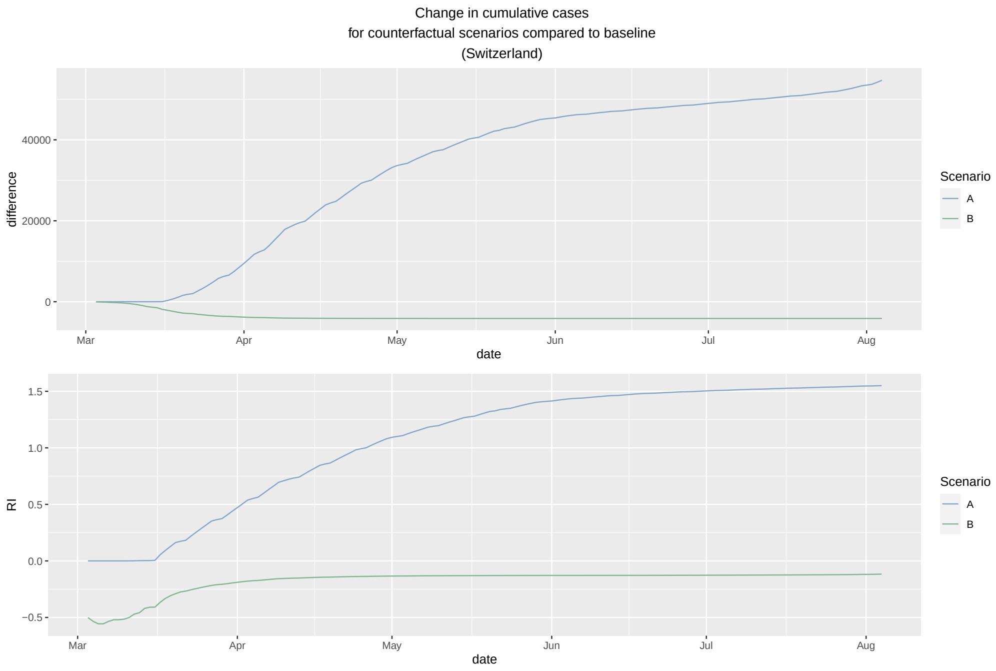


Figure 5: Change in cumulative cases compared to baseline in Switzerland between 2020-03-02 and 2020-08-04

### 3. Comparison of cases predicted under baseline scenario with observed cases

To illustrate prediction accuracy, we compare the daily incidence predicted under the baseline scenario, where every model parameter and time-dependent matrix is set to reality, with observed cases over the study period. The comparisons can be seen below in Figure 6.

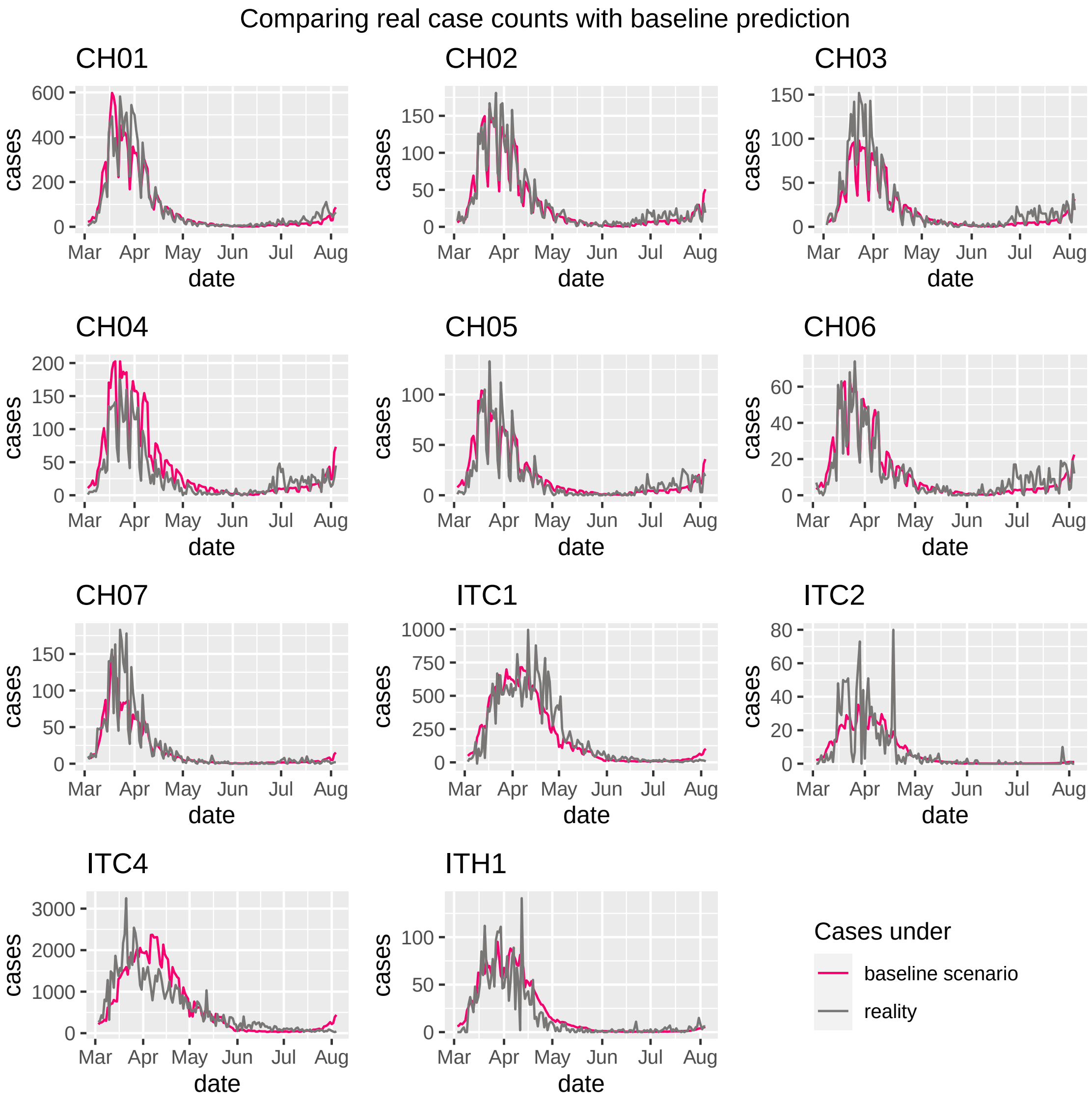


Figure 6: Observed cases and cases predicted under baseline scenario

### 4. Lag distribution

In this section, we plot the estimated lag $u_l$ distribution, that is, the weight each past day is attributed in the autoregressive process. The distribution is seen in Figure 7.

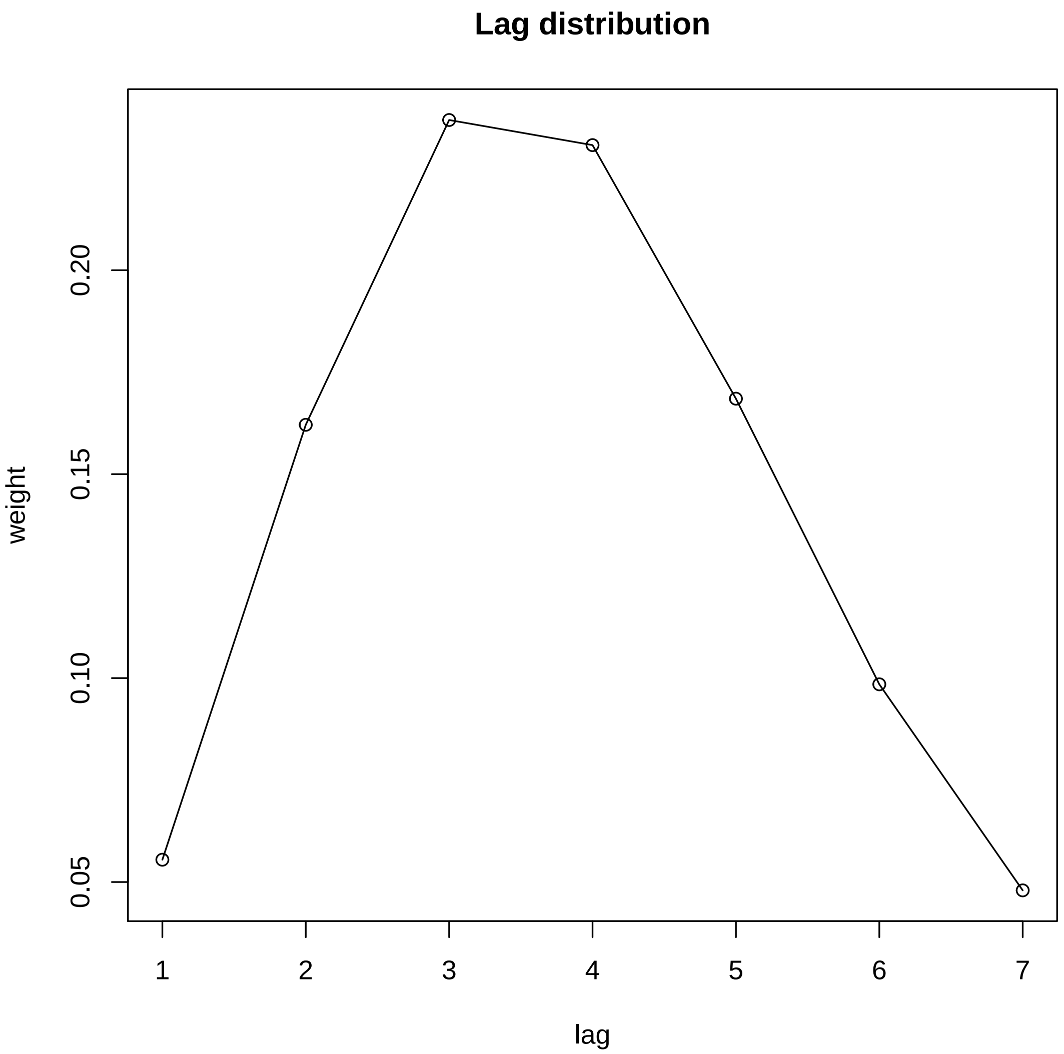


Figure 7: Lag distribution

The lag distribution in our model representing the serial interval corresponds with the 3 to 5 days found by Nishiura et al. [16]. It is also in line with the findings of Ssentongo et al. [17].

[1] Bundesamt für Gesundheit. Covid-19 Schweiz: Informationen zur aktuellen Lage, 2020. URL https://www.covid19.admin.ch/de/api/data.

[2] Presidenza del Consiglio dei Ministri - Dipartimento della Protezione Civile. dati-province, 2020. URL https://github.com/pcm-dpc/COVID-19/blob/master/dati-province/dpc-covid19-ita-province.csv.

[3] Eurostat. Population on 1 January by broad age group, sex and NUTS 3 region (demo_ r_ pjanaggr3), 2021. URL https://ec.europa.eu/eurostat/databrowser/view/demo_r_pjanaggr3/default/table?lang=en.

[4] Swiss Federal Statistical Office. Ständige und nichtständige wohnbevölkerung nach institutionellen gliederungen, geburtsort und staatsangehörigkeit", 2019. URL https://www.pxweb.bfs.admin.ch/pxweb/de/px-x-0102020000_201.

[5] Istituto Nationale di Statistica. Statistiche demografiche ISTAT, 2018. URL http://demo.istat.it/bilmens2019gen/index.html.

[6] Eidgenössisches Justiz- und Polizeidepartement EJPD. Gesetzliche feiertage und tage, die in der schweiz wie gesetzliche feiertage behandelt werden, 2020. URL bj.admin.ch.

[7] Governo Italiano Presidenza del Consiglio dei Ministri. Festività e giornate nazionali, 2020. URL http://demo.istat.it/bilmens2019gen/index.html.

[8] Bundesamt für Gesundheit BAG. New coronavirus: Situation in Switzerland: Conducted tests, 2020. URL https://covid-19-schweiz.bagapps.ch/de-3.html.

[9] European Centre for Disease Prevention and Control. Download data on testing for COVID-19 by week and country, 2020. URL https://www.ecdc.europa.eu/en/publications-data/covid-19-testing.

[10] European Climate Assessment and Dataset. Daily data, 2020. URL https://www.ecad.eu/dailydata/index.php.

[11] International Organization for Migration. Travel restrictions matrix, 2020. URL https://migration.iom.int.

[12] Facebook. Facebook data for good. movement range data, 2020. URL https://data.humdata.org/dataset/movement-range-maps.

[13] Facebook Data for Good. Protecting privacy in Facebook mobility data during the COVID-19 response, 2020. URL https://research.fb.com/blog/2020/06/protecting-privacy-in-facebook-mobility-data-during-the-covid-19-response.

[14] Eurostat. Geodata administartive units/statistical unites, 2016. URL https://ec.europa.eu/eurostat/web/gisco/geodata/reference-data/administrative-units-statistical-units/nuts.

[15] S. Meyer and L. Held. Power-law models for infectious disease spread. *Ann. Appl. Stat*, 8:1612–1639, 2014. doi: 10.1214/14-AOAS743.

[16] H. Nishiura, N. M. Linton, and A. R. Akhmetzhanov. Serial interval of novel coronavirus (COVID-19) infections. *Int. J. Infect. Dis*, 93:284–286, 2020. doi: 10.1016/j.ijid.2020.02.060.

[17] P. Ssentongo, C. Fronterre, A. Geronimo, S. J. Greybush, P. K. Mbabazi, J. Muvawala, S. B. Nahalamba, P. O. Omadi, B. T. Opar, S. A. Sinnar, Y. Wang, A. J. Whalen, L. Held, C. Jewell, A. J. B. Muwanguzi, H. Greatrex, M. M. Norton, P. Diggle, and S. J. Schiff. Tracking and predicting the African COVID-19 pandemic. *medRxiv*, 2020. doi: 10.1101/2020.11.13.20231241.
